# Impact of online health information-seeking behavior on shared decision-making in patients with systemic lupus erythematosus: the TRUMP^2^-SLE project

**DOI:** 10.1101/2023.02.15.23285964

**Authors:** Takanori Ichikawa, Dai Kishida, Yasuhiro Shimojima, Nobuyuki Yajima, Nao Oguro, Ryusuke Yoshimi, Natsuki Sakurai, Chiharu Hidekawa, Ken-ei Sada, Yoshia Miyawaki, Keigo Hayashi, Kenta Shidahara, Yuichi Ishikawa, Yoshiki Sekijima, Noriaki Kurita

## Abstract

**Objective:** Providing appropriate health information to patients with systemic lupus erythematosus (SLE) is advantageous in the treatment decision-making process. We aimed to investigate how online health information-seeking behavior affects shared decision-making (SDM) in patients with SLE.

**Methods:** This cross-sectional study included 464 patients with SLE from five institutions. The main exposure was time spent on the Internet per day, which was divided into four categories (none, <1 h, 1–<2 h, and ≥2 h). Participants categorized their preferred first source of health information as physicians, the Internet, or other media. The outcome was the degree of SDM measured via the 9-item Shared Decision-Making Questionnaire (SDM-Q-9). A general linear model was applied.

**Results:** Compared with no Internet use, longer internet use was associated with a higher SDM-Q-9 score: <1 h, 6.9 points (95% confidence interval [CI] 0.32 to 13.6) and ≥2 h, 8.75 points, (95% CI 0.61 to 16.9). The SDM-Q-9 did not differ between those who chose physicians and those who chose the Internet as their preferred first source of health information (-2.1 points [95% CI -6.7 to 2.6]). Those who chose other media had significantly lower SDM-Q-9 scores than those who chose physicians (-7.6 points [95% CI -13.2 to -1.9]).

**Conclusions:** The present study suggests that SDM between physicians and patients is positively (rather than adversely) associated with online information-seeking behavior, with no negative influence associated with accessing the Internet prior to visits to a rheumatologist. Rheumatologists may need to introduce their patients to websites offering high-quality health information to establish a productive physician–patient relationship for SDM.

## 1. INTRODUCTION

Shared decision-making (SDM), an overarching principle for managing systemic lupus erythematosus (SLE), [1] is a decision-making process in which both patients and medical providers participate in the choice of treatment. [2] Increased efficacy of self-management and satisfaction with treatment regimens ensured by SDM [3,4] lead to the expectation that SDM will contribute to achieving treatment goals among patients with SLE who need to revisit disease-modifying therapies at various life stages. An increasing number of patients seek health information prior to their outpatient visits, and their participation in SDM requires knowledge and understanding of their disease conditions and treatment options. [5,6] Up to 80% of patients with SLE use the Internet to gather information to understand flare-ups, changes in symptoms, and laboratory results. [7] Despite the high frequency of online health information-seeking behavior, its impact on SDM has not been well-documented among patients with SLE. While online information-seeking behavior is expected to improve the physician–patient relationship by serving as an additional resource for patients to better understand physician advice, [8] some studies report that patients do not share information gathered via the Internet with their physicians, and that this behavior can lead to decreased trust in physicians as a result of poor SDM. [6,9] In addition, Internet sources may not have accurate or understandable information, [5,10] and different sources of health information may negatively affect patients’ understanding, resulting in the risk of poor SDM. Recently, as well as 20 years ago, the Internet has been reported to be the second most preferred source of health information after medical professionals. [11,12] However, some patients seek health information from other media sources. Therefore, we aimed to examine how the preference of health information sources and frequency of Internet use affect SDM using data from the Trust Measurement for Physicians and Patients with SLE (TRUMP^2^-SLE) study involving Japanese patients with SLE. Clarifying this will facilitate the understanding of the best sources of information for patients with SLE who will likely make multiple treatment choices with their physicians over their life course.

## 2. METHODS

### 2.1 Study design and participants

This cross-sectional study used baseline data from the TRUMP^2^-SLE study, which is an ongoing multicenter cohort study conducted at five academic medical centers (Showa University Hospital, Okayama University Hospital, Shinshu University Hospital, Yokohama City University Hospital, and Yokohama City University Medical Center).

The inclusion criteria were as follows: (1) patients with SLE aged ≥20 years diagnosed according to the revised 1997 American College of Rheumatology classification criteria, (2) those receiving rheumatology care at the participating center, and (3) those able to respond to the questionnaire survey. Patients with dementia or total blindness were excluded from this study.

### 2.2 Main exposures

1. Time spent on the Internet: The main exposure was time spent on the Internet per day, excluding the time spent working. It was measured by asking, “How much time do you spend on the Internet and social network services in a day, excluding the time spent working?” Respondents were then asked to choose from six choices ranging from “never” to “more than 4 h.” During analysis, the responses were merged into the following categories: not at all, <1 h, 1-<2 h, and ≥2 h.
2. First preference of source of health information: Patients were asked to select the healthcare information source they would like to access first, and responses were categorized as follows: physicians, the Internet, and other media sources (e.g., family and friends, healthcare professionals other than physicians, or TV and radio). Participants were asked a question in Japanese that translates to “Imagine that you are trying to obtain information about your disease and its treatment. Where would you choose to obtain this information from first?”

### 2.3 Outcome measures

The main outcome was SDM, measured using the 9-item Shared Decision-Making Questionnaire (SDM-Q-9; Japanese version). [13–15] The SDM-Q-9 is used by the patient to gauge the level of SDM with their doctors and is valid for use in clinical settings in Japan. [14] The SDM-Q-9 is composed of nine questions scored on a 6-point scale. Patients were asked to choose one of the following responses for each item: ‘completely disagree’ (0 points) to ‘completely agree’ (5 points). The sum of the scores was converted into a scale ranging from 0 to 100. Cronbach’s alpha coefficient for the Japanese version of the SDM-Q-9 was 0.917, demonstrating construct validity.

### 2.4 Measurement of covariates

Confounding variables were those suspected to influence both internet use and SDM based on evidence in the literature. [12] Confounding variables included age, sex, highest educational level, household income, marital status, history of cancer, disease activity, and duration of illness. Disease activity was measured by the attending physician using the Systemic Lupus Erythematosus Disease Activity Index of 2000. The questionnaire was administered at each facility between June 2020 and August 2021, and the patients were asked to complete it either in the waiting room or at home. The questionnaire included assurances that the attending physician would not view the responses, and that answers would only be used at the central facility for aggregation.

### 2.5 Statistical analyses

All statistical analyses were performed using Stata/SE, version 16.1 (StataCorp, College Station, TX, USA). Patient characteristics are described as frequencies and proportions for categorical variables and means and standard deviations (SDs) for continuous variables. The association of the SDM-Q-9 responses with the time spent on the Internet and the first preference for accessing health information was analyzed using a general linear model. The eight patient characteristics described above were included in the multivariate analysis as covariates. In addition, using the general linear model, the SDM-Q-9 scores by Internet usage time and preferences for accessing health information were evaluated via Stata’s margin command and were graphically presented. [16] Missing covariates were addressed using a multiple completion approach. Twenty imputations were performed using multiple imputations with chained equations, assuming that the analyzed data were missing at random. Statistical significance was set at P < 0.05.

### 2.6 Ethical considerations

This study was approved by the ethics committee of Shinshu University Hospital (approval number 5433). Informed consent was obtained from all the participants before enrolment. All the study procedures were performed in accordance with the Declaration of Helsinki and Health Research Involving Human Subjects in Japan. Patient information was anonymized and de-identified prior to analyses.

### 2.7 Patient and public involvement

Neither the general public nor patients with SLE were involved in the planning, recruitment, or conduct of this study.

## 3. RESULTS

### 3.1 Patient flowchart

Initially, 518 patients with SLE who met the inclusion criteria were identified. Among them, 54 patients who did not disclose their duration of Internet use and SDM-Q-9 scores, in addition to patients with duplicate responses for health information source preferences, were excluded. In total, 464 patients were included in the analysis (Fig. 1).

**Figure 1.**
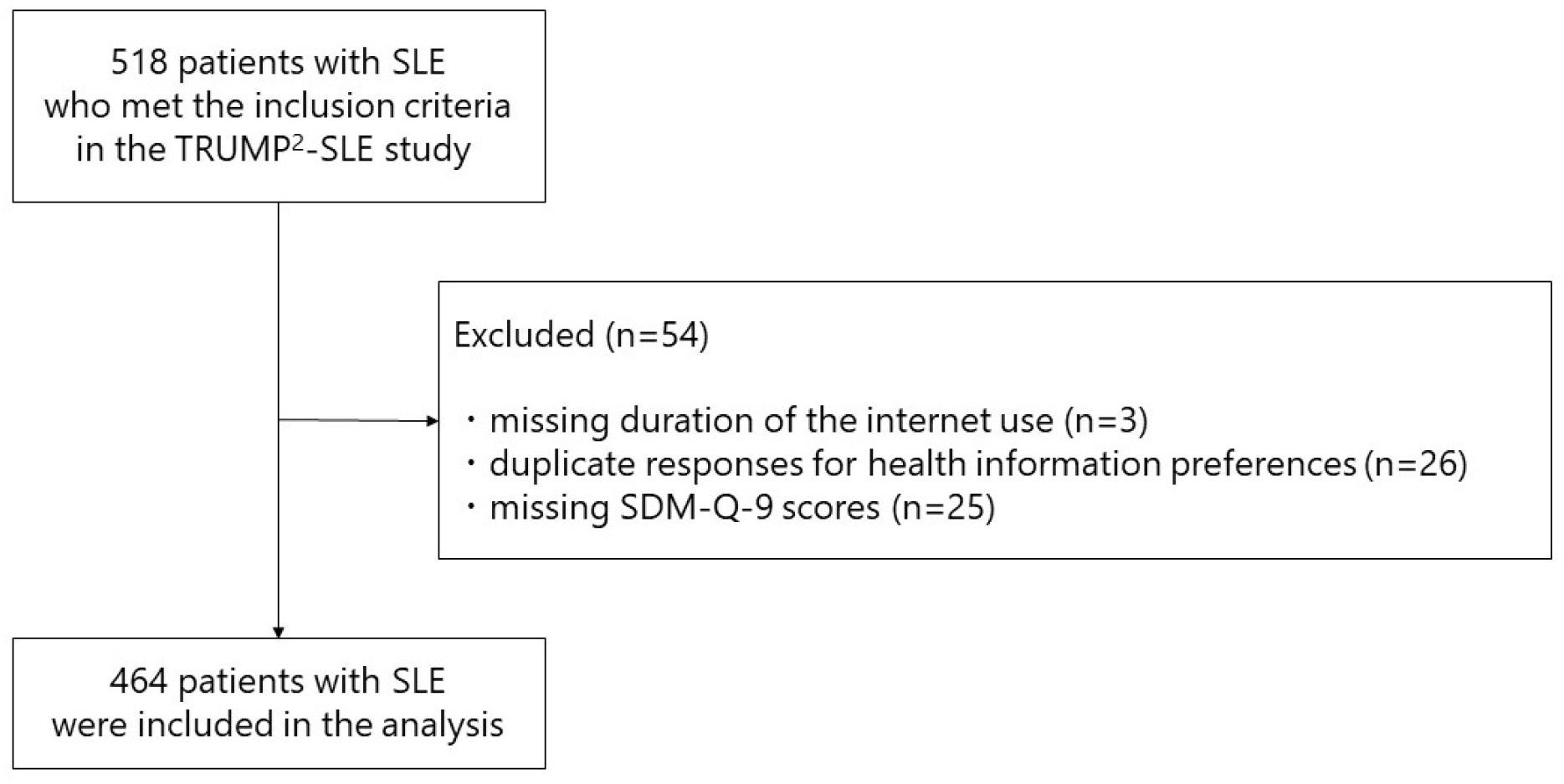
Patient flowchart. SDM-Q-9 score, 9-item Shared Decision-Making Questionnaire; SLE, systemic lupus erythematosus.

### 3.2 Patient characteristics

Patient characteristics by Internet use are shown in Table 1.

**Table 1.**
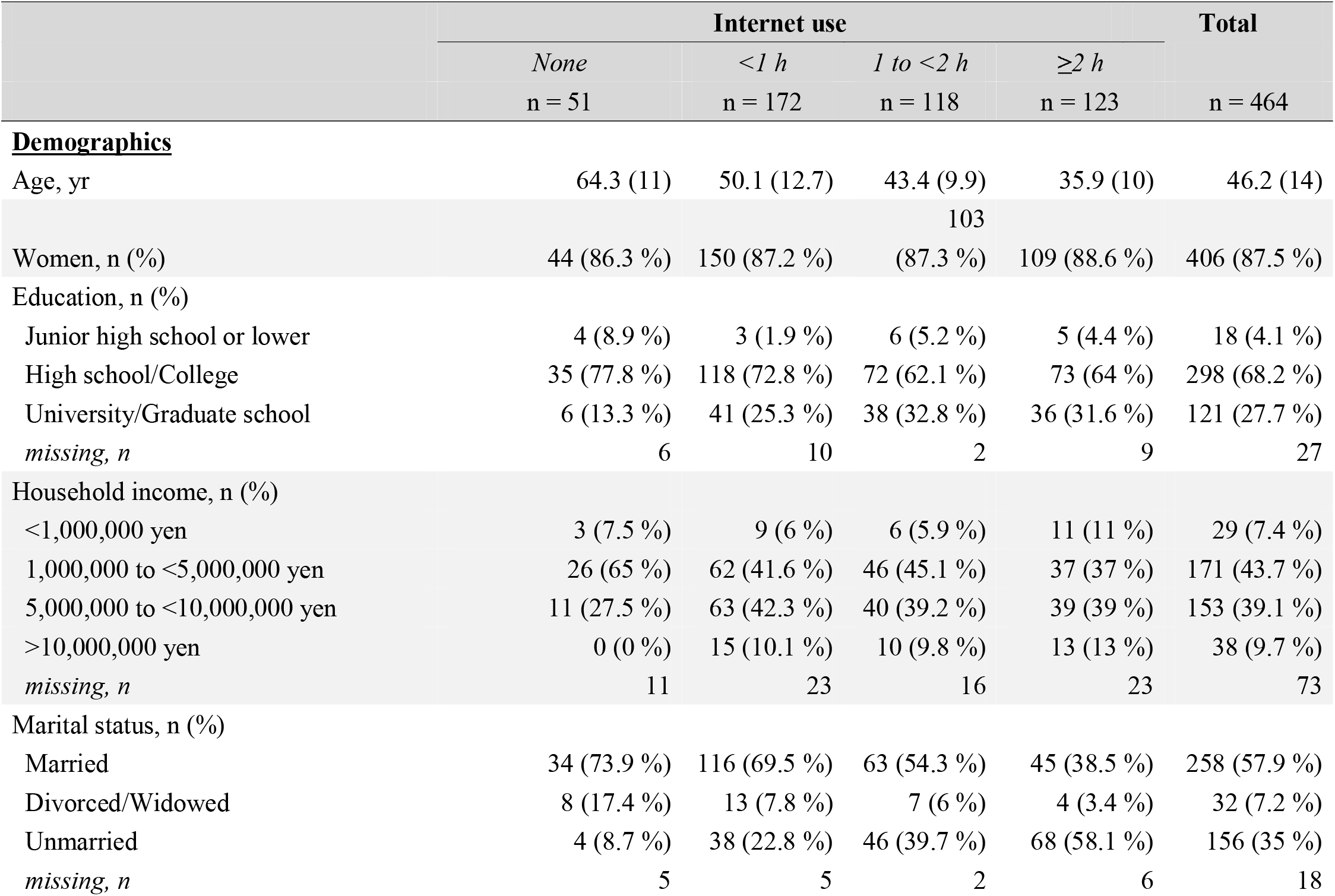
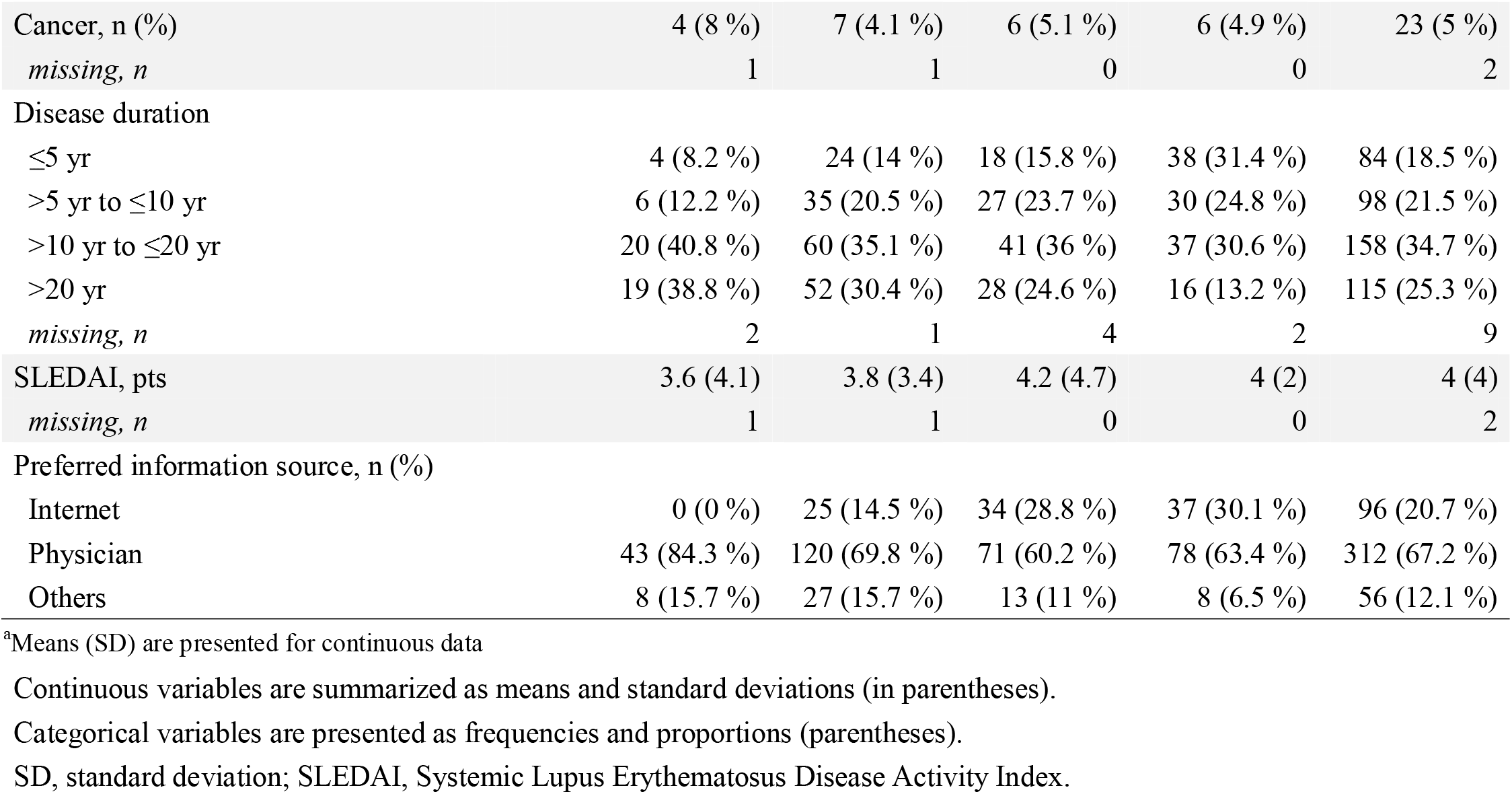
Characteristics of patients separated by Internet use (n = 464)

The mean age of the patients was 46 years (SD, 14 years), and 406 patients (88%) were women. Participants who reported longer Internet use (≥2 h/day) were younger, more likely to be unmarried (58.1%), had a shorter disease duration (≤5 years, 31.4%), and more likely to have completed university/graduate school (31.6%). Moreover, those who reported longer Internet use per day were more likely to favor it as their primary source of health information.

### 3.3 Patient characteristics by the first preference for accessing health information

Patient characteristics based on the preference for accessing health information are shown in the Supplementary Table. Patients who preferred the Internet were younger and more likely to have a disease duration ≤10 years.

### 3.4 Internet use, first preference for accessing health information, and SDM

The relationships between Internet use duration, first preference for accessing health information, and SDM are shown in Table 2. Longer Internet use tended to be associated with a higher SDM-Q-9 score than no Internet use (<1 h use, 6.97 points, 95% confidence interval [CI], 0.32 to 13.6; 1 to <2 h use, 7.05 points, 95% CI -0.38 to 14.5; ≥2 h use, 8.75 points, 95% CI 0.61 to 16.9). The SDM-Q-9 score did not differ among those who preferred the Internet for obtaining health information and those who preferred their attending physicians (-2.1 points, 95% CI -6.7 to 2.6). In contrast, those who preferred other sources of information had a significantly lower SDM-Q-9 (-7.6 points, 95% CI -13.2 to -1.9).

**Table 2.**
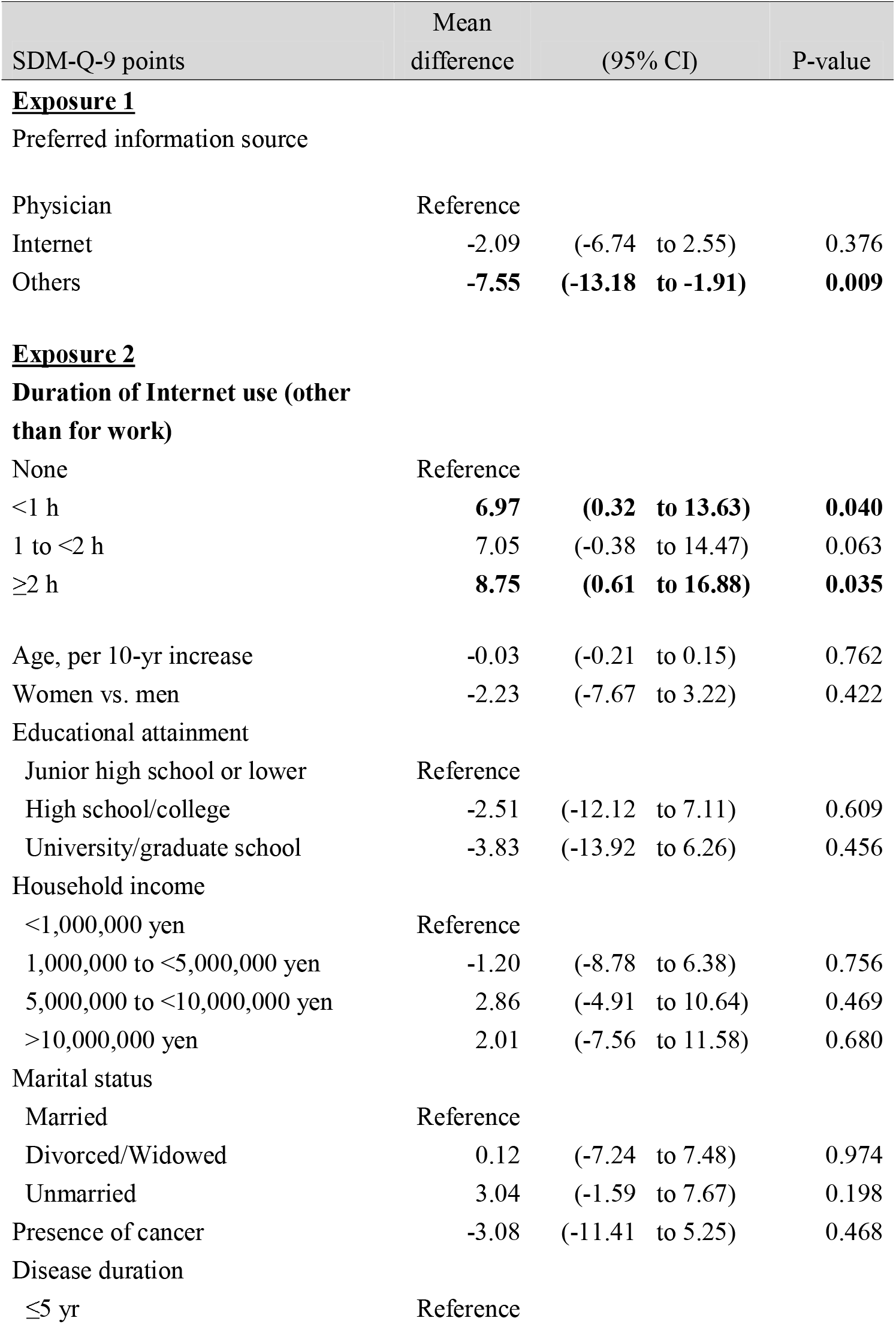
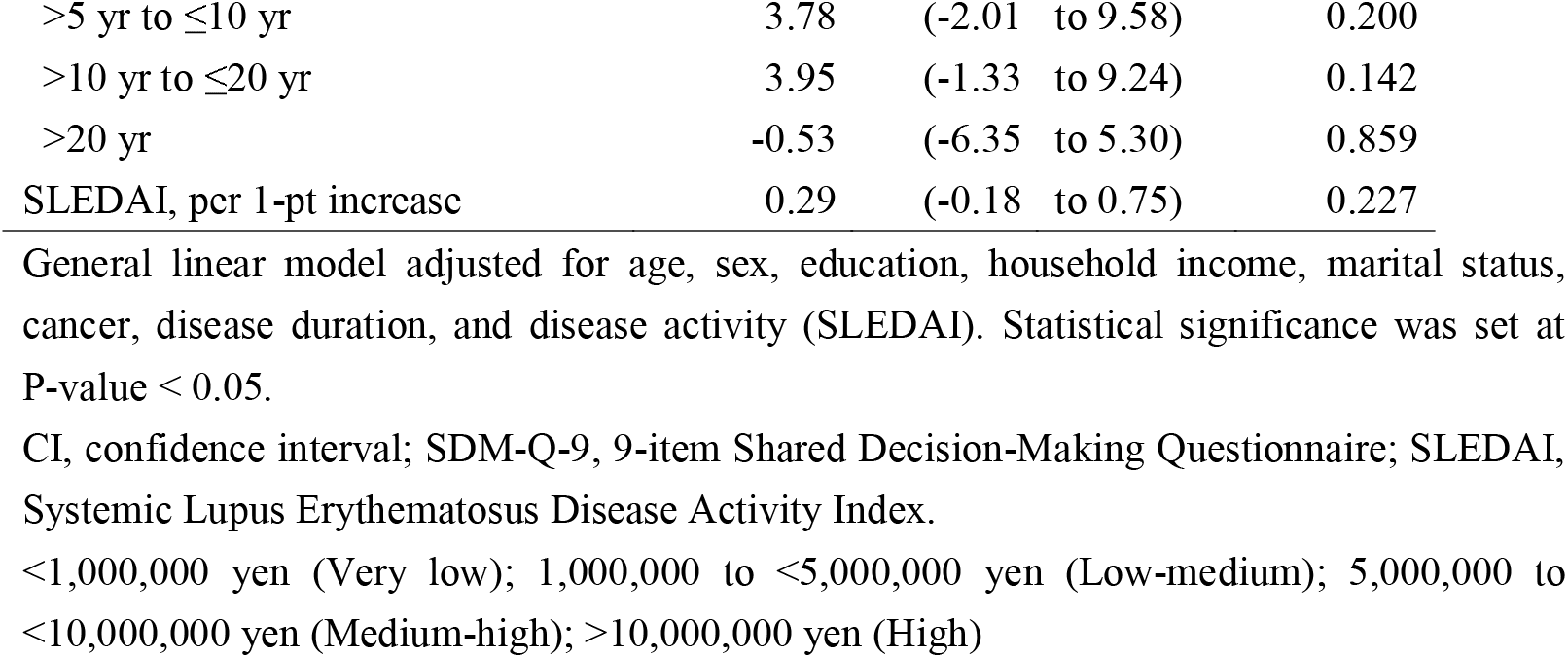
Association of shared decision-making with preferred information source and duration of Internet use† (n = 464)

| SDM-Q-9 points | Mean difference | (95% CI) | P-value |
| --- | --- | --- | --- |
| **<u>Exposure 1</u>** |  |  |  |
| Preferred information source |  |  |  |
| Physician | Reference |  |  |
| Internet | -2.09 | (-6.74 to 2.55) | 0.376 |
| Others | **-7.55** | **(-13.18 to -1.91)** | **0.009** |
| **<u>Exposure 2</u>** |  |  |  |
| **Duration of Internet use (other than for work)** |  |  |  |
| None | Reference |  |  |
| <1 h | **6.97** | **(0.32 to 13.63)** | **0.040** |
| 1 to <2 h | 7.05 | (-0.38 to 14.47) | 0.063 |
| ≥2 h | **8.75** | **(0.61 to 16.88)** | **0.035** |
| Age, per 10-yr increase | -0.03 | (-0.21 to 0.15) | 0.762 |
| Women vs. men | -2.23 | (-7.67 to 3.22) | 0.422 |
| Educational attainment |  |  |  |
| Junior high school or lower | Reference |  |  |
| High school/college | -2.51 | (-12.12 to 7.11) | 0.609 |
| University/graduate school | -3.83 | (-13.92 to 6.26) | 0.456 |
| Household income |  |  |  |
| <1,000,000 yen | Reference |  |  |
| 1,000,000 to <5,000,000 yen | -1.20 | (-8.78 to 6.38) | 0.756 |
| 5,000,000 to <10,000,000 yen | 2.86 | (-4.91 to 10.64) | 0.469 |
| >10,000,000 yen | 2.01 | (-7.56 to 11.58) | 0.680 |
| Marital status |  |  |  |
| Married | Reference |  |  |
| Divorced/Widowed | 0.12 | (-7.24 to 7.48) | 0.974 |
| Unmarried | 3.04 | (-1.59 to 7.67) | 0.198 |
| Presence of cancer | -3.08 | (-11.41 to 5.25) | 0.468 |
| Disease duration |  |  |  |
| ≤5 yr | Reference |  |  |
| >5 yr to ≤10 yr | 3.78 | (-2.01 to 9.58) | 0.200 |
| >10 yr to ≤20 yr | 3.95 | (-1.33 to 9.24) | 0.142 |
| >20 yr | -0.53 | (-6.35 to 5.30) | 0.859 |
| SLEDAI, per 1-pt increase | 0.29 | (-0.18 to 0.75) | 0.227 |

General linear model adjusted for age, sex, education, household income, marital status, cancer, disease duration, and disease activity (SLEDAI). Statistical significance was set at P-value < 0.05.
CI, confidence interval; SDM-Q-9, 9-item Shared Decision-Making Questionnaire; SLEDAI, Systemic Lupus Erythematosus Disease Activity Index.
<1,000,000 yen (Very low); 1,000,000 to <5,000,000 yen (Low-medium); 5,000,000 to <10,000,000 yen (Medium-high); >10,000,000 yen (High)

### 3.5 SDM-Q-9 scores predicted by time spent on the Internet per day and preferred information source

The predicted SDM-Q-9 scores based on the time spent on the Internet per day and preferred information source are shown in Fig. 2. The SDM-Q-9 score was higher in participants with a longer Internet use duration. The SDM-Q-9 score was 66.9 with no Internet use [60.6 to 73.2] and 75.7 with >2 h of Internet use [71.7 to 79.6]. Preferring to obtain health information from physicians resulted in an SDM-Q-9 of 74.9 points [72.8 to 77.1], whereas “other” information sources resulted in an SDM-Q-9 of 67.4 points [62.2 to 72.6].

**Fig 2.**
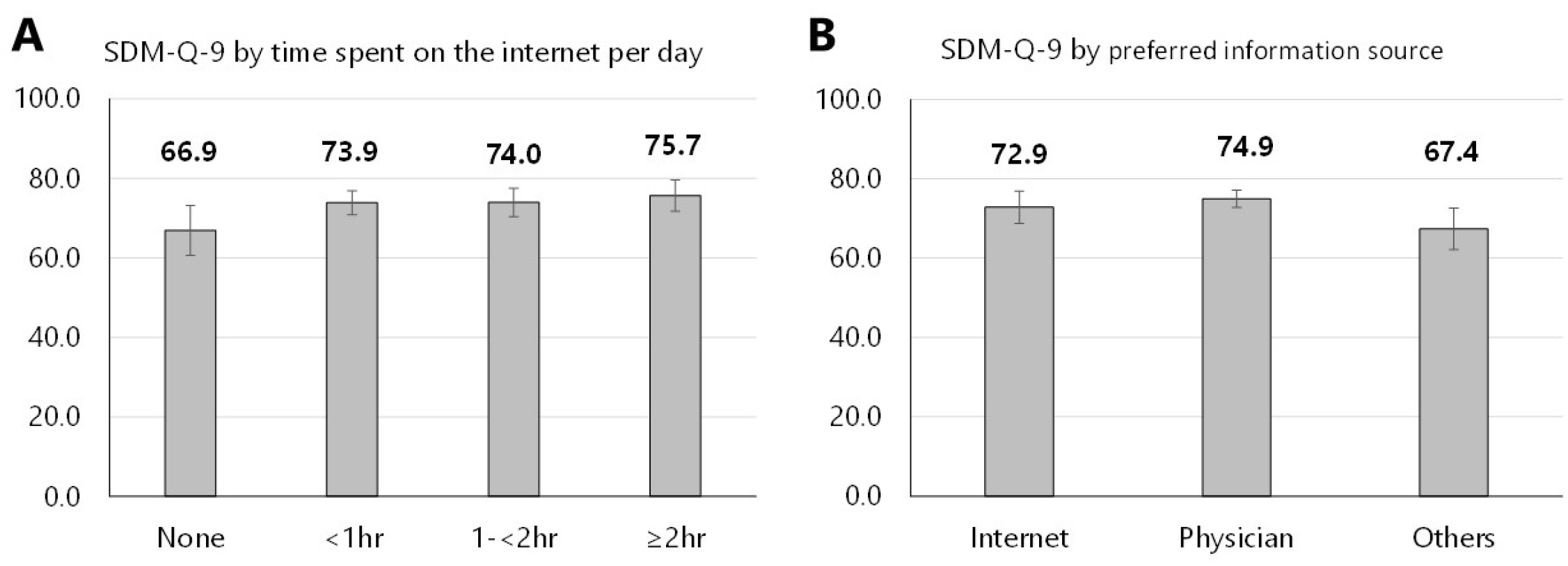
SDM-Q-9 scores predicted by time spent on the internet per day and preferred information source (n=464). A: Predicted mean scores on the SDM-Q-9 calculated by the time spent on the internet per day. B: Predicted mean scores on the SDM-Q-9 calculated by preferred information source. SDM-Q-9 scores were predicted by the time spent on the Internet or by the preferred source of health information. The adjusted SDM-Q-9 scores were predicted using the general linear model described in Table 2. The left vertical axis indicates the adjusted SDM-Q-9 scores. Error bars indicate 95% confidence intervals. Gray bars indicate point estimates. For example, the predicted mean SDM-Q-9 score would be 66.9 (95% confidence interval 60.6–73.2) if the entire analyzed population reported no Internet use. SDM-Q-9, nine-item Shared Decision-Making Questionnaire

## 4. DISCUSSION

### 4.1 Short summary

We examined the influence of the duration of online information-seeking behavior and first preference for health information sources on SDM among patients with SLE. The extent of SDM was positively correlated with Internet use time. The extent of SDM did not differ between those who chose a physician and those who chose the Internet as their first choice for accessing information.

### 4.2 Comparison with the results of previous studies

The findings of the present study are consistent with those of previous studies on the factors influencing SDM in patients with SLE. Specifically, previously identified factors associated with SDM among patients with SLE were predominantly physician–patient interactions, such as physician empowerment and trust in physicians.[3,17] To our knowledge, the present study is the first to include the Internet when exploring patients’ health information-seeking behavior.

### 4.3 Explanations

The association between longer Internet use and better SDM was contrary to our hypothesis that longer Internet use would hinder SDM because patients exposed to more uncertain information would be more skeptical of their physicians’ suggestions. While a possible association between online health information-seeking behavior and dissatisfaction with the provision of information by physicians has been noted, some patients with SLE believe that online health information is complementary to that provided by their physicians. [18] In addition to the complementary benefits of online health information-seeking behavior, a previous finding found that sharing online health information with physicians led to higher satisfaction with office visits for both patients and physicians, further supporting our findings. [6,18] A younger age, higher education level, and shorter disease duration among the patients with longer Internet use observed in the present study are consistent with those described in previous studies. [18,19] However, the relationship between longer Internet use and better SDM, independent of age, education, and disease duration, is noteworthy. Reportedly, 62.5% of patients with rheumatic diseases gather health information online before their first visit. [6] Consequently, the impact of online information-seeking behavior, which is common, on SDM between physicians and patients cannot be underestimated. The rationale for the high SDM-Q-9 scores obtained by patients who preferred the Internet as their first source of health information, as well as by those who preferred physicians, can be explained by considering that pre-visit information seeking serves as a pre-learning process, facilitating physicians and patients to fully discuss treatment options together. [5] Even if patients are exposed to a massive amount of health information, some of which may be biased and problematic, [6] an open discussion with physicians could help reduce uncertainty about the disease course and the optimal treatment plan. These conversations steer SDM toward the best choice for each patient. A previous study reinforced this notion, showing that primary care patients who use the Internet ask more questions during office visits and thus follow physicians’ recommendations more rigorously. [20] Our finding that the proportion of patients who prefer the Internet as their first choice for accessing health information is higher with a longer Internet use duration is consistent with the findings of a Spanish study of patients with SLE, in which online health information seeking was more prevalent among regular Internet users. [18]

### 4.4 Clinical implications

This study has several implications for clinicians working with patients with SLE. First, asking patients with SLE how much they use the Internet or whether they search for health information online may provide an opportunity for rheumatologists to facilitate SDM. Considering that patients do not always share the health information they find on the Internet with their physicians [6,18] and do not want to damage their relationship with their physicians by sharing it, [6] such an inquiry would provide insights into the patient’s health information-seeking behavior and foster frank dialogue. Second, because patients have difficulty assessing the accuracy of online information [5] and interpreting its relevance to their illness, [21] physicians can use the opportunity to link their expertise with patients’ knowledge by helping them interpret online health information [18] or by referring them to high-quality websites. [18] Finally, efforts to provide high-quality online information to patients who do not use the Internet are needed. Patients who did not use the Internet were older and had a longer disease duration. Strategies could include providing patients with tablet-based treatment decision aids in an outpatient setting or encouraging the patients’ relatives or friends to help them find information online. [18]

### 4.5 Strengths

The present study has several strengths. To our knowledge, this is the first study to demonstrate the impact of patients’ preferences for health information sources and Internet use duration as factors influencing the degree of SDM among patients with SLE, after adjusting for confounders such as age, disease activity, education level, and economic status. Second, the findings come from several centers, including those in metropolitan and rural areas, and may be generalizable to a broader population. Third, this study is timely and important from a socio-medical standpoint because we were able to examine the effects of online information-seeking behavior on SDM in an era when online media, such as websites and social network services, has become widespread.

### 4.6 Limitations

The present study had some limitations. First, reverse causation is possible because of the nature of cross-sectional studies. For example, insufficient engagement in SDM may result in patients preferring access to other media to supplement their knowledge. Second, the strength of the correlation between the time spent on the Internet and time spent gathering health information among the patients is unknown. However, considering the results of a previous study that showed a greater frequency of online health information seeking among patients with SLE who were regular Internet users, [18] we believe that time spent on the Internet can approximate the time spent online gathering health information.

## 5. CONCLUSIONS

In conclusion, this study demonstrated that online health information-seeking behavior is positively associated with SDM among patients with SLE. Therefore, rheumatologists should consider incorporating innovative strategies to facilitate patients’ access to reliable online health information in their clinical practice and proactively communicate with patients based on this information to help them make better decisions regarding health management plans.

## Data Availability

All data produced in the present study are available upon reasonable request to the authors.

## Abbreviations

CI: confidence interval
SD: standard deviations
SDM: shared decision-making
SDM-Q-9: 9-item Shared Decision-Making Questionnaire
SLE: systemic lupus erythematosus
SLEDAI: Systemic Lupus Erythematosus Disease Activity Index
TRUMP^2^-SLE: Trust Measurement for Physicians and Patients with SLE

## 6. ACKNOWLEDGEMENTS

We would like to thank Hiroko Nagasato, Kumi Sasaki, Yukari Hosaka (Showa University), and Miyuki Sato (Fukushima Medical University) for their support. The abbreviated name of the study, “TRUMP^2^-SLE (the Trust Measurement for Physicians and Patients with SLE),” does not refer to a specific individual; instead, the name intends to suggest a trusted physician, as an old meaning of the word “trump” is “a dependable and exemplary person.”

## COMPETING INTERESTS

NK is a member of the Committee on Clinical Research, Japan College of Rheumatology, and has received grants from the Japan Society for the Promotion of Science, consulting fees from GlaxoSmithKline K.K., and payments for speaking and educational events from Chugai Pharmaceutical Co. Ltd., Sanofi K.K., Mitsubishi Tanabe Pharma Corporation, and Japan College of Rheumatology. K. Sada has received a research grant from Pfizer Inc. and a payment for speaking and educational events from GlaxoSmithKline K.K. The remaining authors declare no competing interests.

## FUNDING

This study was supported by JSPS KAKENHI (Grant Number: JP 19KT0021). The funder had no role in the study design, analyses, interpretation of the data, writing of the manuscript, or decision to submit the manuscript for publication.

## DATA AVAILABILITY STATEMENT

The datasets generated and/or analyzed during the study are available from the corresponding author upon reasonable request.

## Notes

### Author Declarations

This study was approved by the ethics committee of Shinshu University Hospital (approval number 5433).

