## Supplementary Table 1 for "Impact of online health information-seeking behavior on shared decision-making in patients with systemic lupus erythematosus: the TRUMP^2^-SLE project"

**Supplementary Table 1. Characteristics of patients by preferred information source (n= 464)**

|  | **preferred information source** | | | **Total** |
| --- | --- | --- | --- | --- |
|  | *Internet* | *Physician* | *Others* |  |
|  | n = 96 | n = 312 | n = 56 | n = 464 |
| **Demographics** |  |  |  |  |
| Age, yr | 41.9 (12.3) | 46.5 (14) | 52 (14.8) | 46.2 (14) |
| Women, n (%) | 86 (89.6 %) | 274 (87.8 %) | 46 (82.1 %) | 406 (87.5 %) |
| Education, n (%) |  |  |  |  |
| Junior high school or lower | 6 (6.5 %) | 10 (3.5 %) | 2 (3.7 %) | 18 (4.1 %) |
| High school/college | 63 (67.7 %) | 196 (67.6 %) | 39 (72.2 %) | 298 (68.2 %) |
| University/graduate school | 24 (25.8 %) | 84 (29 %) | 13 (24.1 %) | 121 (27.7 %) |
| *missing, n* | 3 | 22 | 2 | 27 |
| Household income, n (%) |  |  |  |  |
| <1,000,000 yen | 8 (9.8 %) | 18 (6.8 %) | 3 (6.7 %) | 29 (7.4 %) |
| 1,000,000 to <5,000,000 yen | 37 (45.1 %) | 116 (43.9 %) | 18 (40 %) | 171 (43.7 %) |
| 5,000,000 to <10,000,000 yen | 32 (39 %) | 103 (39 %) | 18 (40 %) | 153 (39.1 %) |
| >10,000,000 yen | 5 (6.1 %) | 27 (10.2 %) | 6 (13.3 %) | 38 (9.7 %) |
| *missing, n* | 14 | 48 | 11 | 73 |
| Marital status, n (%) |  |  |  |  |
| Married | 49 (52.7 %) | 173 (57.9 %) | 36 (66.7 %) | 258 (57.9 %) |
| Divorced/Widowed | 9 (9.7 %) | 20 (6.7 %) | 3 (5.6 %) | 32 (7.2 %) |
| Unmarried | 35 (37.6 %) | 106 (35.5 %) | 15 (27.8 %) | 156 (35 %) |
| *missing, n* | 3 | 13 | 2 | 18 |
| Cancer, n (%) | 2 (3.1 %) | 17 (7.4 %) | 4 (10.3 %) | 23 (6.9 %) |
| *missing, n* | 0 | 2 | 0 | 2 |
| Disease duration |  |  |  |  |
| ≤5 yr | 16 (16.8 %) | 60 (19.5 %) | 8 (15.1 %) | 84 (18.5 %) |
| >5 yr to ≤10 yr | 25 (26.3 %) | 67 (21.8 %) | 6 (11.3 %) | 98 (21.5 %) |
| >10 yr to ≤20 yr | 33 (34.7 %) | 103 (33.6 %) | 22 (41.5 %) | 158 (34.7 %) |
| >20 yr | 21 (22.1 %) | 77 (25.1 %) | 17 (32.1 %) | 115 (25.3 %) |
| *missing, n* | 1 | 5 | 3 | 9 |
| SLEDAI, pts | 3.6 (3.3) | 4.2 (4.3) | 3.3 (3) | 4 (4) |
| *missing, n* | 0 | 2 | 0 | 2 |
| Duration of Internet use (other than for work), n (%) |  |  |  |  |
| None | 0 (0 %) | 43 (13.8 %) | 8 (14.3 %) | 51 (11 %) |
| <1 h | 25 (26 %) | 120 (38.5 %) | 27 (48.2 %) | 172 (37.1 %) |
| 1 to <2 h | 34 (35.4 %) | 71 (22.8 %) | 13 (23.2 %) | 118 (25.4 %) |
| ≥2 h | 37 (38.5 %) | 78 (25 %) | 8 (14.3 %) | 123 (26.5 %) |

Continuous variables are summarized as means and standard deviations (in parentheses).

Categorical variables are presented as frequencies and proportions (in parentheses).

SLEDAI, Systemic Lupus Erythematosus Disease Activity Index.
